# Determinants of multidrug-resistant urinary tract infections: a retrospective cross-sectional study from a tertiary care hospital in southern Bangladesh

**DOI:** 10.1101/2025.08.01.25332668

**Authors:** Ibrahim Khalil, Abu Sayed, A.K.M. Akbar Kabir, Md. Nurul Alam, S. M. Iqbal Hossain, Rahima Akther Dipa, Md Tanvir Rahman

## Abstract

Multidrug resistance (MDR) poses significant public health concerns with global impacts, yet data on factors driving MDR in urinary tract infections (UTIs) within Bangladesh remains scarce. Therefore, this study aimed to identify factors associated with MDR in UTI patients. A retrospective cross-sectional study was conducted from January to December 2023 at a tertiary care hospital in Barishal, Bangladesh. Among 1,797 urine samples, 229 with significant bacterial growth were analyzed. Antimicrobial susceptibility testing (AST) was performed using the disk diffusion method. Univariate and multivariate logistic regression analyses were applied to determine significant predictors and risk factors. Multiple Correspondence Analysis (MCA) was used to prioritize categories of predictors and risk factors, highlighting primary and secondary contributors to MDR in UTI patients. Out of 229 samples assessed, 60 were MDR-positive. *E. coli* (55.9%) was the predominant uropathogen, followed by *Pseudomonas* spp. (20.5%), *Klebsiella* spp. (14.8%), and *Acinetobacter* spp. (8.7%). Samples from specialized units (SUs) had significantly higher odds of MDR (OR = 5.9; *p* = 0.002) compared to outpatient department (OPD). Additionally, male patients from private medical settings (PMSs) exhibited higher odds (OR = 21.8; *p* = 0.004). MCA identified MDR-positive status and SU sample sources as primary contributors, while *Acinetobacter* spp., and *Pseudomonas* spp. were linked to secondary dimensions. The findings highlight SU samples and male patients from PMSs as key predictors of MDR, underscoring the need for further research and the implementation of evidence-based antimicrobial stewardship (AMS) in resource-limited settings.

## Introduction

Multidrug resistance (MDR), which is a subset of antimicrobial resistance (AMR), impacts human well-being, environmental systems, and economic stability. In contrast to AMR, which includes resistance to all types of antimicrobial agents (such as antibiotics, antivirals, antifungals, and antiparasitics), MDR is the state of a species of microorganism to at least one antimicrobial drug in three or more antimicrobial categories [1]. MDR increases both morbidity and mortality by rendering standard antibiotic treatments ineffective, leading to prolonged illnesses, and higher rates of treatment failure [2]. A study from the World Bank reported that AMR could lower GDP by 2.45% in low-income nations and lead to an alarming global economic shortfall of $16.7 trillion by 2050 (World Bank, 2016) [3]. It also harms ecosystems through contamination via wastewater, agricultural runoff, and improper disposal methods, facilitating horizontal gene transfer that further spreads resistance [4]. In healthcare settings, inanimate surfaces like bedrails, stethoscopes, and supply carts are often contaminated through patient shedding or contact with healthcare workers, facilitating the spread of multidrug-resistant organisms (MDROs) and reducing treatment effectiveness [5–6]. Therefore, World Health Organization (WHO) remarked AMR as one of the top ten global public health hazards, urgently requires coordinated international actions to combat its increasing effects on health, environment, and economic structures [7].

Urinary tract infections (UTIs) are among the most widespread bacterial infections globally, impacting around 150 million people each year in community settings [8]. They also rank as the fifth most frequently occurring healthcare-associated infection (HAI), significantly contributing to worldwide mortality, with 62,700 deaths directly linked to them in 2015 [9]. The main causative agent of UTIs is uropathogenic *Escherichia coli* (UPEC), followed by pathogens such as *Klebsiella pneumoniae*, *Proteus mirabilis*, *Enterococcus faecalis*, and various *Staphylococcus* spp. [10]. The prevalence of uropathogens varies depending on age, sex, and other factors, with notably higher infection rates in females and significant association seen in individuals aged 19 years old or older [11–12]. Standard treatment for UTIs typically involves antibiotic classes like trimethoprim-sulfamethoxazole, fluoroquinolones, nitrofurantoin, fosfomycin, and β-lactams, with the success of treatment hinging on achieving effective antimicrobial levels in the urine [13–15]. Nevertheless, the growing occurrence of MDR uropathogens poses a serious challenge to effective treatment. Alarmingly, high rates of MDR have been recorded among *E. Coli* (77.6%), *Pseudomonas* spp. (90.5%), *Acinetobacter baumannii* (88.5%), and *Klebsiella* spp. (7.3-100%) [16–18]. The high frequency of MDR pathogens highlights the pressing need for thorough surveillance, improved AMS, and the exploration of alternative treatment options to address this global health issue.

MDR-UTIs result from a complicated interaction of demographic, clinical, and environmental elements, creating a substantial challenge for healthcare systems worldwide. Age and sex are significant predisposing factors, as individuals over 65 years and males show higher prevalence rates [19–20]. Other associated risk factors for MDR-UTIs include prior antibiotic use, recent hospitalization, urinary catheterization, nursing home residence, diabetes mellitus, and recurrent infections [21–23]. The clinical origin of urine samples can influence MDR trends; Shakya *et al.* (2021) reported a higher frequency of MDR isolates in samples obtained from inpatient departments (e.g., medicine, surgery, nephrology, obstetrics) compared to outpatient clinics, with rates of 66.2% and 49.5%, respectively [24]. Warm weather may be associated with the occurrence of MDR, as rising temperatures can affect bacterial behavior and promote the development of antibiotic resistance patterns [25].

Despite the well-documented global impact of MDR, there remains a critical gap in understanding the interplay of demographic, clinical, and environmental factors contributing to resistance, especially in low- and middle-income countries (LMICs). In the south-west part of Bangladesh, no study has been conducted to identify the factors associated with MDR, which is crucial for local AMS programs. Moreover, the MDR pattern is alarmingly changing over time and across locations, hindering targeted initiatives [26]. Therefore, this research was undertaken with the aim to determine the predictors and risk factors associated with MDR in UTIs in south-western part of Bangladesh and provide evidence-based insights to inform AMS programs tailored to LMICs. The findings have far-reaching implications, offering a model for similar investigations in resource-constrained settings and contributing to the global fight against antimicrobial resistance.

## Methods

### Study location

This study was conducted at the Microbiology Laboratory of Sher-E-Bangla Medical College & Hospital (SBMCH), Barishal, a government-operated tertiary care hospital of south-western territory of Bangladesh (**Fig 1**).

**Fig. 1.**
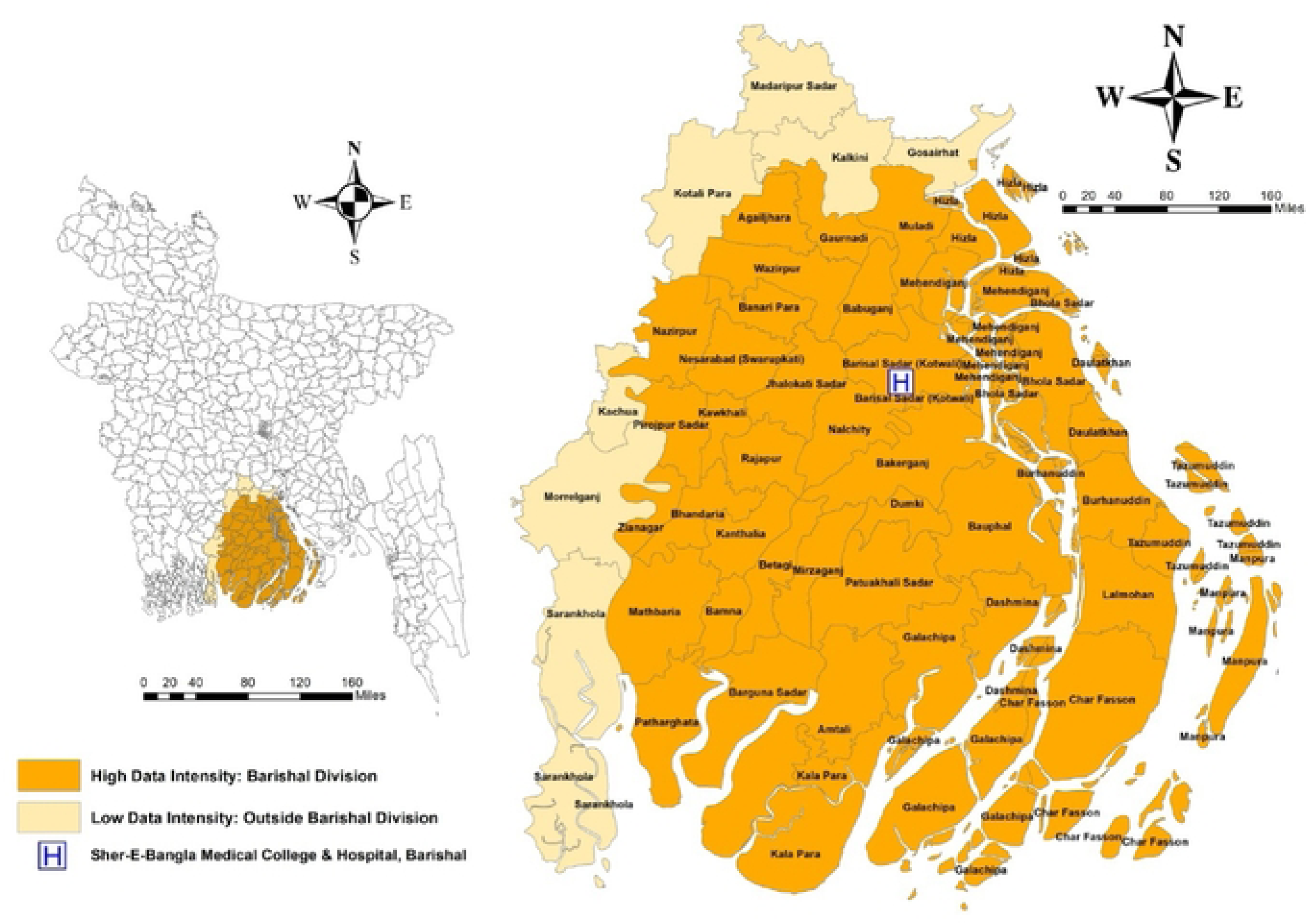
Geographical Distribution of obtained Urine Samples from UTI Patients across Barishal Division and Surrounding Areas, Bangladesh

### Study design and duration

A retrospective cross-sectional study was conducted using an anonymized secondary dataset comprising urine culture and sensitivity test results collected between January and December 2023 at Sher-E-Bangla Medical College & Hospital (SBMCH), Barishal, a government-operated tertiary medical care facility in southwestern Bangladesh. A total of 1,797 urine samples were initially collected from patients suspected of UTIs as part of routine clinical diagnostics. Among these, 1,420 samples originated from the outpatient department (OPD), 180 from specialized units (SUs), 70 from private medical settings (PMSs), and 127 lacked source specification in the dataset.

The present study did not involve direct contact with patients or any additional sample collection beyond routine clinical procedures. As the dataset was fully anonymized prior to analysis, individual informed consent was not applicable. A waiver of consent and approval for the secondary use of clinical data were granted by the Institutional Review Board (IRB) of SBMC.

### Sample collection and preparation

At first, midstream, clean-catch urine samples were obtained from each patient using wide-mouth, screw-capped, sterile containers, adhering to standard precautions to prevent contamination. Urine samples, each with a minimum volume of 5 mL, were collected from each patient and transported to the microbiology laboratory within 30 minutes of collection. The samples were inoculated into Sheep Blood Agar and MacConkey Agar media and then incubated aerobically at 37 °C for 16–18 hours [27]. Bacterial growth was considered significant if the colony count was ≥10⁵ CFU/mL of a typical urinary tract infection [28].

### Laboratory analysis

#### Bacterial isolation and identification

Cultures exhibiting significant bacterial growth were first assessed using Gram staining technique to classify the isolates as Gram-positive or Gram-negative domain. Gram-negative bacteria were identified using a series of biochemical tests, including motility, urease, and indole tests in MIU (Motility-Indole-Urea) medium; Triple Sugar Iron (TSI) agar for carbohydrate utilization; citrate utilization; and oxidase tests. Gram-positive bacteria were confirmed through catalase and coagulase tests, ensuring precise identification of bacterial species by observing their cultural patterns.

#### Antimicrobial Susceptibility Testing (AST)

Bacterial colonies were transferred to Tryptic Soy Broth and incubated at 37°C to achieve a turbidity equivalent to the 0.5 McFarland standards, ensuring standardized inoculum density. A sterile cotton swab was dipped into the prepared suspension, excess fluid was removed, and the swab was used to uniformly streak the surface of Mueller-Hinton Agar plates in three directions. Antibiotic discs were placed on the agar surface using a sterile dispenser, and the plates were incubated at 37°C for 16–18 hours. Zones of inhibition were measured and interpreted according to the Clinical Laboratory Standards Institute (CLSI) guidelines, 2022. AST was performed using a broad range of antibiotics, categorized into several groups, including Aminoglycosides (Amikacin, Gentamycin, Tobramycin, Netilmycin), Beta-lactam/Beta-lactamase inhibitor combinations (Amoxicillin-Clavulanate, Piperacillin-Tazobactam), Penicillins (Ampicillin, Penicillin), Cephalosporins (Cefazolin, Cefepime, Cefexime, Cefotaxime, Cefoxitin, Ceftaroline, Ceftazidim-Avibactam, Ceftazidime, Ceftriaxone, Cefuroxime), Carbapenems (Imipenem, Meropenem), Fluoroquinolones (Ciprofloxacin, Levofloxacin, Nalidixic acid), Macrolides (Azithromycin, Erythromycin), Tetracyclines (Doxycycline, Tetracycline, Tigecycline), Glycopeptides (Vancomycin), Sulfonamides (Sulfamethoxazole-Trimethoprim), Phenicols (Chloramphenicol), Lincosamides (Clindamycin), Polypeptides (Colistin), Fosfomycin, Trimethoprim, and Oxazolidinones (Linezolid). However, due to limitations in the availability of certain antibiotic discs, uniform drug sensitivity testing could not be performed for all the isolates.

### Data cleaning and management

Data from laboratory findings and patient records were entered into an Excel spreadsheet and carefully cleaned to ensure accuracy and completeness. Only urine samples with single bacterial colonies were considered for analysis (229 out of a total of 1797 samples). Antibiotic names were standardized and categorized into their respective antibiotic classes for determination of MDR. Variables were eventually recoded and then coded for further statistical analysis.

### Data analysis and visualization

#### Descriptive analysis

Descriptive analysis was performed to determine the proportions and 95% confidence intervals (CIs) of demographic, seasonal, and microbial variables, including sex, age, season of sample collection, sample source, and bacterial growth. Proportions were calculated as percentages of the total sample size (N=229) and summarized in tabular format for clear presentation.

#### Univariate analysis

The study assessed the univariate association between multidrug-resistant (MDR) status and various predictors and risk factors. Chi-square tests were used to evaluate the significance of differences in proportions between MDR-positive and MDR-negative cases for categorical variables. Fisher’s exact test was applied where expected cell frequencies were less than 5. The variables analyzed included Season (categorized as Winter, Spring, Summer, and Autumn, where Winter includes December, January, and February; Spring encompasses March, April, and May; Summer covers June, July, and August; and Autumn comprises September, October, and November), Age Group (1–18 years [Child], 19–35 years [Young Adult], 36–55 years [Middle-aged Adult], 56–74 years [Older Adult], and 75+ years [Elderly]), Sex (Female, Male), Source of Sample (OPD, PMSs, and SUs), with SUs comprising special units within Sher-E-Bangla Medical College such as the Female Surgery Unit, Male Surgery Unit, Pediatric Unit, Labor Unit, Medicine Unit, Female Medicine Unit, Urology Unit, and other departments dedicated to specialized care, and Bacterial Growth (*Acinetobacter* spp.*, E. coli, Klebsiella* spp.*, and Pseudomonas* spp.). Statistical significance was set at *p* < 0.05. Results were presented as percentages and counts of MDR-positive and MDR-negative cases, with Pearson Chi-square values used to assess the strength of associations.

#### Multivariate analysis

A multivariate logistic regression model was developed to identify independent and confounding predictors and risk factors for MDR-UTIs. Adjusted odds ratios with 95% CIs were calculated to quantify the degree of association between predictors and risk factors and MDR status. Interaction terms were included to evaluate effect modification across key predictors, and confounding was assessed by comparing unadjusted and adjusted ORs. Model evaluation was possessed using Hosmer-Lemeshow test to assess goodness-of-fit, followed by sensitivity, specificity, positive predictive value (PPV), and negative predictive value (NPV) to evaluate classification performance. Predicted probabilities for MDR-positive classification were generated, and the default classification threshold of 0.5 was adjusted to 0.4 and 0.3 to improve sensitivity. Receiver operating characteristic (ROC) analysis was conducted to visualize model performance, and the area under the ROC curve (AUC) was calculated to measure discriminatory power. Multicollinearity among factors was assessed using variance inflation factors (VIFs). Additionally, the Net Reclassification Index (NRI) and Integrated Discrimination Improvement (IDI) were calculated to evaluate improvements in classification and discrimination after the inclusion of interaction terms.

#### Multiple Correspondence Analysis (MCA)

Multiple Correspondence Analysis (MCA) was employed to examine the associations and underlying structure among categorical variables potentially associated with multidrug-resistant urinary tract infections (MDR-UTIs). MCA is a dimensionality reduction technique specifically designed for categorical data, allowing for the visualization and interpretation of complex relationships between multiple variables.

In this study, each categorical variable was first transformed into a series of binary (dummy) variables. A Burt matrix—a symmetrical matrix summarizing all pairwise cross-tabulations of categories—was then constructed. To extract the principal dimensions, Singular Value Decomposition (SVD) was applied to the Burt matrix. This process decomposes the data into orthogonal dimensions (or components), with each dimension capturing a proportion of the total inertia (variance) in the dataset.

The first dimension (Dimension 1) explains the greatest proportion of the total variance, followed by subsequent dimensions (e.g., Dimension 2) capturing progressively less. The contribution of each category to these dimensions was quantified by evaluating their coordinates along the respective axes, weighted by their inertia. Categories with higher contributions are considered more influential in defining the structure and relationships within the dataset.

Eigenvalues derived from the SVD process were used to determine the proportion of total variance explained by each dimension. Dimensions explaining a cumulative variance of over 60– 70% were considered sufficient to capture the major patterns within the data. The resulting graphical and numerical outputs facilitated the identification of clusters and associations between patient characteristics, sample sources, bacterial isolates, and MDR status. All the statistical analyses were performed using the Statistical Software for Data Science-STATA (version 13, StataCorp, USA).

#### Geographic mapping

The figures for this study were created using Canva, and the study map was developed using Geographic Information System software (ArcGIS, version 10.8, Esri, Redlands, CA, USA).

### Ethical considerations

Ethical approval for this study was obtained from the Institutional Review Board (IRB) of SBMC, affiliated with the University of Dhaka, under memo number 59.14.0000.130.99.001.24.2763, dated 28 November 2024. The study involved the retrospective analysis of fully anonymized secondary data obtained from laboratory records of urine samples submitted for diagnostic purposes.

As the data were de-identified and no interventions or direct patient interactions occurred, the IRB granted a waiver of informed consent. All procedures adhered to the institutional ethical guidelines and complied with the principles outlined in the Declaration of Helsinki.

## Results

### Demographic, seasonal, and isolated bacterial profile

The majority of samples were from females (71.2%), young adults (36.2%), autumn (30.1%), and the OPD (65.5%), compared to males, other age groups, seasons, and sample sources. *E. coli* was the predominant bacterial isolate (55.9%), followed by *Pseudomonas* spp. (20.5%), *Klebsiella* spp. (14.8%), and *Acinetobacter* spp. (8.7%) **Table 1**.

**Table 1.** Demographics, Seasonality, and Bacterial Isolates of Urine Samples from UTI Patients in 2023 (N = 229)

| Variable | Category | % (n) | 95% CI |
| --- | --- | --- | --- |
| Sex | Female | 71.2 (163) | 0.65, 0.77 |
|  | Male | 28.8 (66) | 0.23, 0.34 |
| Age (Years) | Child (1–18) | 10.9 (25) | 0.06, 0.15 |
|  | Young Adult (19–35) | 36.2 (83) | 0.30, 0.42 |
|  | Middle-aged Adult (36–55) | 24.0 (55) | 0.18, 0.29 |
|  | Older Adult (56–74) | 20.1 (46) | 0.14, 0.25 |
|  | Seniors (75+) | 8.7 (20) | 0.05, 0.12 |
| Season | Autumn | 30.1 (69) | 0.24, 0.36 |
|  | Spring | 27.1 (62) | 0.21, 0.32 |
|  | Summer | 21.8 (50) | 0.16, 0.27 |
|  | Winter | 21.0 (48) | 0.15, 0.26 |
| Sample Source | OPD | 65.5 (150) | 0.59, 0.71 |
|  | PMSs | 17.9 (41) | 0.13, 0.23 |
|  | SUs | 16.6 (38) | 0.12, 0.21 |
| Bacterial Isolates | *E. coli* | 55.9 (128) | 0.49, 0.62 |
|  | – OPD | 70.3 (90) |  |
|  | – PMSs | 14.8 (19) |  |
|  | – SUs | 14.8 (19) |  |
|  | *Pseudomonas* spp. | 20.5 (47) | 0.15, 0.25 |
|  | – OPD | 51.1 (24) |  |
|  | – PMSs | 21.3 (10) |  |
|  | – SUs | 27.6 (13) |  |
|  | *Klebsiella* spp. | 14.8 (34) | 0.10, 0.19 |
|  | – OPD | 61.8 (21) |  |
|  | – PMSs | 23.5 (8) |  |
|  | – SUs | 14.7 (5) |  |
|  | *Acinetobacter* spp. | 8.7 (20) | 0.05, 0.12 |
|  | – OPD | 75.0 (15) |  |
|  | – PMSs | 20.0 (4) |  |
|  | – SUs | 5.0 (1) |  |

Outpatient Department-OPD; Specialized Units-SUs; Private Medical Settings-PMSs

### Univariate association

Significant associations were observed between MDR prevalence and age, sex, and source of sample. The Elderly (75+ years) group had the highest MDR prevalence (50%), followed by the Older Adult (56-74 years) group (45.7%). Males exhibited a significantly higher MDR rate (50%) compared to females (22.7%) (**Fig 2**). Among the sample sources, specialized units had the highest MDR prevalence (57.9%), followed by private practice (46.3%), while OPD samples showed the lowest MDR rate (19.3%).

**Fig. 2:**
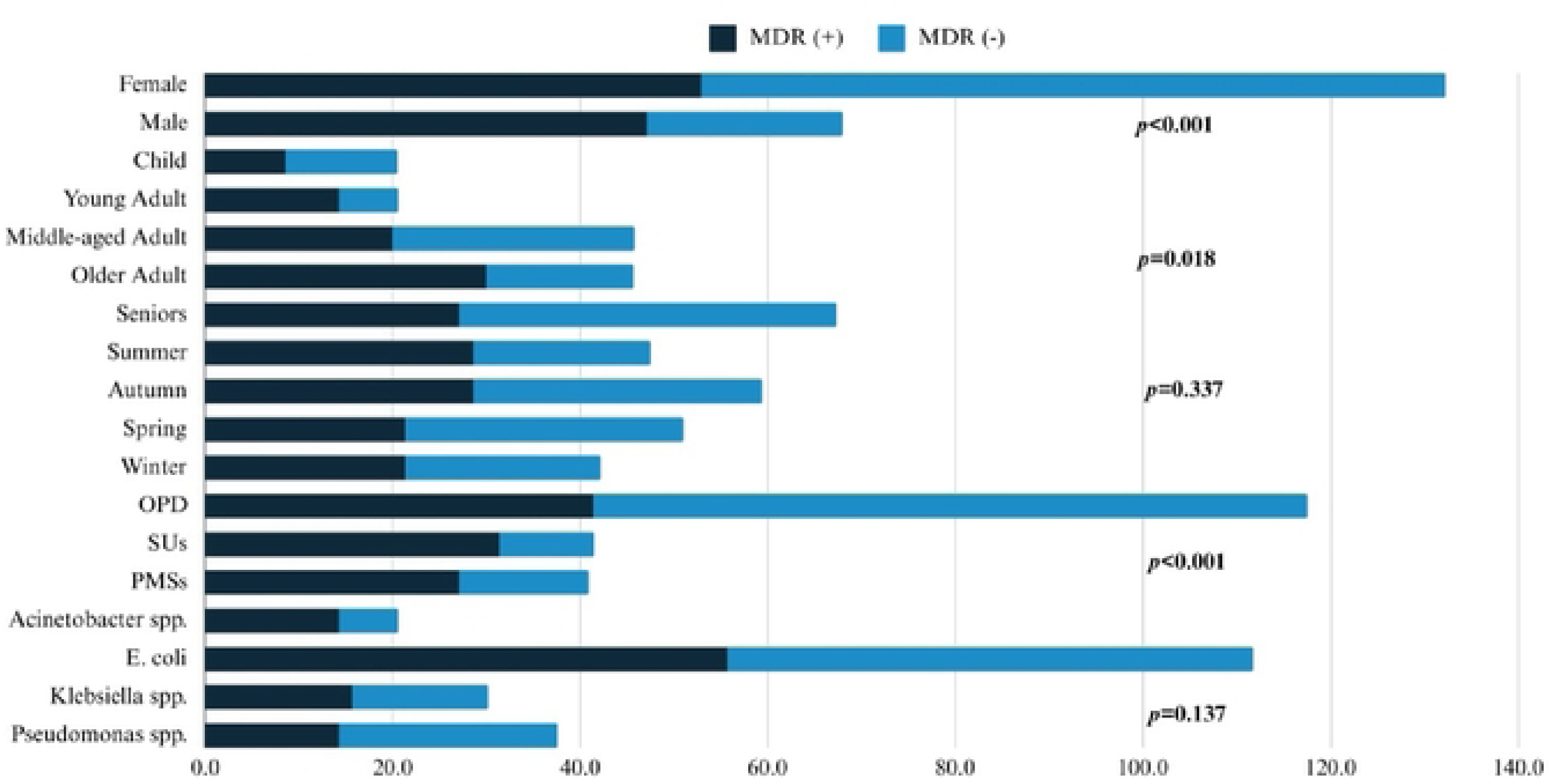
Univariate Analysis of MDR Positivity across Categories

### Multivariate relationship

Samples from SUs had significantly higher odds of MDR, in contrast to OPD (OR = 5.9; *p* = 0.002). Among bacterial isolates, *E. coli* (OR = 0.3; *p* = 0.022) and *Pseudomonas* spp. (OR = 0.1; *p* = 0.000) were found to have significantly lower odds of MDR when compared to *Acinetobacter* spp. Furthermore, males from PMSs had markedly higher ratios for MDR (OR = 21.8; *p* = 0.004), indicating a critical subgroup for intervention **Table 2**.

**Table 2:**
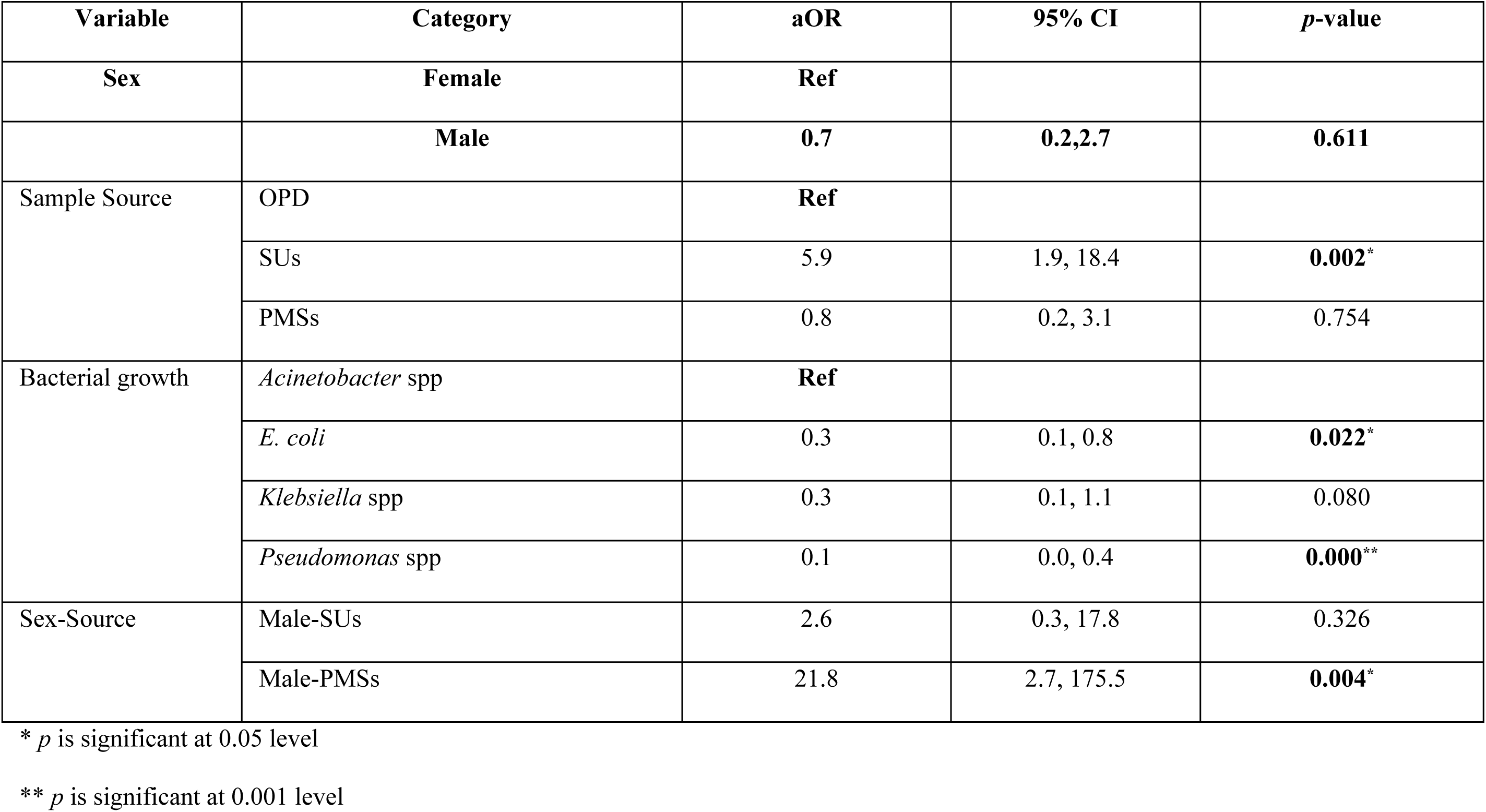
Multivariate Logistic Regression Analysis (MLRA) of Factors Associated with Multidrug-Resistant (MDR) Status.

The Hosmer-Lemeshow test demonstrated a good model fit (*p* = 0.96), indicating no significant evidence of poor calibration. At the default threshold of 0.5, the model achieved high specificity (91.82%) while maintaining moderate sensitivity (41.43%), effectively identifying MDR-negative cases without compromising performance. Adjusting the classification threshold to 0.4 increased sensitivity to 55.71%, with a slight reduction in specificity to 85.45%, while further lowering the threshold to 0.3 increased sensitivity to 70.00%, although specificity decreased to 78.00%, reflecting the trade-off between early detection and false positives. The model’s positive predictive value (PPV) was 69.05%, ensuring that 69.05% of predicted MDR-positive cases were true positives, while the negative predictive value (NPV) of 78.07% ensured accuracy in identifying MDR-negative cases. The area under the receiver operating characteristic (ROC)<u>g</u> curve (AUC) was 0.7698, indicating good discriminatory power, typically considered in the range of 0.7 to 0.8. The inclusion of the interaction term between Sex and Source significantly improved model performance, as confirmed by a likelihood ratio test (χ² = 8.55, *p* = 0.0139). The Net Reclassification Improvement (NRI) of 0.27 (*p* = 0.010) and the Integrated Discrimination Improvement (IDI) of 0.058 (*p* = 0.004) further suggest significant improvements in risk classification and model discrimination compared to baseline. Multicollinearity was assessed using variance inflation factors (VIFs), with all values below 3.3, which is well below the threshold typically considered problematic (VIF > 5 or 10), suggesting no multicollinearity concerns. These findings highlight the model’s potential for clinical application, with tailored threshold selection to balance sensitivity and specificity while maintaining good discrimination and reliability.

### Multiple Correspondence Analysis (MCA)

To explore the complex interrelationships among categorical predictors associated with multidrug-resistant urinary tract infections (MDR-UTIs), a MCA was performed. The analysis extracted two primary dimensions that jointly explained 78.65% of the total variability in the dataset—Dimension 1 accounted for 75.81%, while Dimension 2 explained an additional 2.84%.

Dimension 1 captured the principal structure of the data and was primarily shaped by categories such as MDR-positive status (Coordinate: −1.464) and samples collected from SUs (Coordinate: −1.950), both of which exhibited strong contributions to this axis. This indicates a strong association between MDR status and certain clinical sources of infection. Conversely, MDR-negative cases (Coordinate: 0.644) and samples from OPD (Coordinate: 0.898) were positioned on the opposite side of this dimension, reflecting contrasting patterns of distribution.

Dimension 2, although accounting for a smaller proportion of variability, highlighted the contribution of specific bacterial isolates. Notably, *Acinetobacter* spp. (Coordinate: 3.928) and *Pseudomonas* spp. (Coordinate: −2.426) exhibited substantial influence on this secondary axis, suggesting their distinct roles in the development of MDR-UTIs.

The MCA plot **Table 3** provided a visual representation of the proximities and relationships among the different categories. Variables contributing more prominently to the dimensions are considered more influential in determining the overall data structure. These findings underscore critical associations between sample sources, bacterial species, and MDR status, which could inform more targeted surveillance and intervention strategies to combat antimicrobial resistance in clinical UTI cases.

**Table 3:** Contribution and Coordinates of Categories in MCA for MDR status in UTI patients.

| Variable | Category |  | Overall |  | Dimension 1<br>(Primary) |  | Dimension 2<br>(Secondary) |  |
| --- | --- | --- | --- | --- | --- | --- | --- | --- |
|  |  | Mass | Quality | % Inertia | Coordinate | Contribution | Coordinate | Contribution |
| Sex | Female | 0.178 | 0.848 | 0.089 | 0.747 | 0.099 | 0.1 | 0.002 |
|  | Male | 0.072 | 0.848 | 0.219 | **-1.844** | **0.245** | -0.248 | **0.004** |
| Sample<br>Source | OPD | 0.164 | 0.801 | 0.125 | 0.898 | 0.132 | 0.081 | 0.001 |
|  | PMSs | 0.045 | 0.778 | 0.098 | -1.479 | 0.098 | **1.162** | **0.06** |
|  | SUs | 0.041 | 0.776 | **0.158** | **-1.95** | **0.158** | **-1.572** | **0.103** |
| **Bacterial growth** | *Acinetobacter* spp. | 0.022 | 0.371 | 0.026 | -0.041 | 0 | **3.928** | **0.337** |
|  | *E. coli* | 0.14 | 0.854 | 0.008 | 0.249 | 0.009 | 0.014 | 0 |
|  | *Klebsiella* spp. | 0.037 | 0.182 | 0.006 | 0.026 | 0 | 0.99 | 0.036 |
|  | *Pseudomonas* spp. | 0.051 | 0.564 | 0.047 | **-0.68** | **0.024** | **-2.426** | **0.302** |
| **MDR** | Negative | 0.174 | 0.813 | 0.069 | **0.644** | 0.072 | **-0.522** | **0.047** |
|  | Positive | 0.076 | 0.813 | **0.156** | **-1.464** | **0.164** | **1.185** | **0.107** |

This table presents the mass, quality, inertia, coordinates, and contributions of variables across two dimensions. Mass indicates the relative weight of each category, while quality reflects the accuracy of representation in the two dimensions. % Inertia shows the variance explained by each dimension. Coordinates represent the positioning of each category along the primary (Dimension 1) and secondary (Dimension 2) axes. Contribution values indicate the relative importance of each category in defining the respective dimensions. Dimension 1 accounts for 75.81% of the total inertia, Dimension 2 accounts for 2.84%, resulting in a cumulative variance of 78.65%, and Dimension 3 explains 0.34% only. Total inertia of the dimensions was 70.93%.

## Discussion

Antimicrobial stewardship (AMS) poses significant challenges particularly in the developing countries due to complexities in governing the health sector [29]. Inadequate health facilities, lack of strict regulations and awareness hinder the AMR, exaggerating the prevalence of MDR organisms [30]. Therefore, our study concluded the critical insights for comprehending the significant predictors and risk factors for MDR patterns in UTI patients from a tertiary care hospital.

In our study, females (71.2%) were found to be more affected by UTIs than males, which are consistent with various studies [31]. For example, a study of Aveiro, Portugal reported almost 80% females to have UTIs, whereas this figure was 20.4% in male patients [32]. Additionally, we found a significantly higher prevalence of MDR-positive cases in females (52.9%), compared to males. This is due to anatomical and physiological factors such as shorter urethras with long diameters and close proximity of the UT to GIT enhance the risk of bacterial translocations, predisposing to have more chances of UTIs than males [33]. Moreover, recurrent infections in females lead to higher frequency of antibiotic uses, ultimately driving the emergence of MDR pathogens [34].

MLRA further demonstrated no significant relationship between the MDR status and sex (OR = 0.7, 95% CI: 0.2–2.7, *p* = 0.611), suggesting other factors such as healthcare exposure, type of organism causing UTIs, playing crucial roles for MDR risk. To cite an example, males who were under PMSs showed the greater odds (OR = 21.8, *p* = 0.004) than SU settings. It clearly indicates the multifactorial involvement of MDR, underscoring the importance of focusing on other predictors and risk factors rather than solely focusing on demographic characteristics of the patients, which is supported by Keenan *et al.,* 2024 [35].

Young adults (36.2%) aged from 19-35 years showed the highest positive cases for UTIs, whereas the infection rate gradually declined in accordance with the age. Interestingly, our study revealed less susceptibility of children to UTIs, aligning with a study carried out in India [36]. However, a number of studies acknowledged the older age as a significant risk factor for UTIs that are not supporting our current results [37–38]. In addition, we noted a positive influence of age (*p* = 0.018) on MDR status of the patients. This positive trend is due to weakened immunity, prolonged exposure to healthcare environment and other senile effects that are linked for the escalation of infection risk, antibiotic uses and ultimately high MDR occurrences. Moreover, longer hospital stays and invasive procedures further increase susceptibility of patients to MDR-UTIs [39]. Despite finding the positive correlation of age with MDR status of patients, no significant confounding effects were observed. Contrary to our findings, Milovanovic with his team had identified age as an independent risk factor for the development of MDR associated UTIs [40].

Previous studies have highlighted seasonal variation as a contributor to UTI prevalence, though, it is not true always [41–42]. Therefore, we considered seasonality as an important factor in relation to UTI occurrences. We observed the highest frequencies in autumn; nevertheless, infection rates across all seasons were almost similar, demonstrating slight variation. In a study of California, Elser and his colleagues identified a clear monotonic inclination in UTI diagnosis in winter when the temperature falls, while the least cases were recorded during summer months [43]. This could be due to increased dehydration and reduced fluid intake in the warmer seasons, resulting in reduction of urine output and subsequently the elimination of uropathogens, act as environmental risk factors for UTIs [44]. Although, seasonal variation was considered as predictor for UTIs, we could not establish any direct impact of climate on development of MDR pathogens, and therefore, could not determine seasonal variation as an independent risk factor.

In terms of samples obtained from different healthcare settings, OPD samples showed the highest prevalence of UTIs. Additionally, sample source was found to be associated with MDR, and samples from SU had significantly higher odds (OR = 5.9; *p* = 0.002), in comparison to the OPD. Moreover, through MLRA, the male sex were observed for MDR-UTIs in PMSs had better odds than in SUs (OR = 21.8; *p* = 0.004). This indicates that the healthcare associated factors are a strong independent predictor for MDR-UTIs in the human patients, supported by a number of studies [45–47].

Among the four (4) bacteria identified from the samples, *E. coli* was the most prevalent (55.9%) among the other gram negative organisms. A comparable study conducted in Iraq identified *E. coli* as the most frequent pathogen (68.3%) which is aligned with our current study finding [48]. Besides that, several other studies have recognized *E. coli* as the paramount contributor for UTIs in human patients which are evident from different healthcare settings throughout the world [49–52].

Our research reveals significant differences in MDR patterns of the bacterial isolates form different clinical settings causing UTIs. *E. coli* (OR = 0.3; *p* = 0.022) and *Pseudomonas spp.* (OR = 0.1; *p* = 0.000) exhibited significantly lower odds than *Acinetobacter* spp., despite no univariate relationship between the bacterial species detected and MDR profile. We used *Acinetobacter* spp. as the baseline for MLR model for comparison because of its high medical significance in relation to MDR and critical healthcare-associated infections, although *E. coli* was detected as the most dominant organism contributing for UTIs in our investigation. These results emphasize the importance of implementing effective control strategies against *Acinetobacter* spp. as a notable and prevalent MDR pathogen, as highlighted by past studies [53–55]. Additionally, the lower odds for *E. coli* and *Pseudomonas* spp. suggest varied resistance mechanisms, underscoring the need for pathogen-specific treatment approaches and further monitor and research for their resistance patterns in order to control the development and spread of resistant genes and organisms through agent-host-environment interphase.

In this study, we conducted MCA for providing a detailed exploration of MDR patterns revealing that Dimension 1 accounts for 75.81% of the total variance, while Dimension 2 contributing an additional 2.84%. Dimension 1 primarily demonstrates the variability driven by healthcare-associated factors, such as the strong association between MDR-positive status and SUs. This finding underscores the critical role of hospital environments in the emergence and propagation of MDR pathogens. In contrast, Dimension 2 highlights the secondary but significant contributions of specific bacterial isolates, with *Acinetobacter* spp. (33.7%) and *Pseudomonas* spp. (30.2%), emerging as dominant contributors. These results are aligned with global reports identifying these pathogens as high-priority targets for infection control [56–60].

### Strengths and Limitations

This study is the first study reported from south-western region of Bangladesh, which employs a robust multifaceted approach to identify independent predictors as well as risk factors for MDR patterns, ensuring deeper insights into healthcare-associated risks and bacterial resistance profiles. It emphasizes pathogen-specific insights, such as resistance patterns in *E. coli*, *Pseudomonas* spp., and *Acinetobacter* spp., aligning with global infection control priorities. Additionally, the incorporation of seasonal variations and alignment with global data enhances the study’s relevance and scope for practical applications, particularly in AMS and targeted infection control strategies.

Although the study was conducted in a tertiary care hospital, its findings may not be entirely pertinent to other healthcare settings, such as rural or primary care facilities. The limited sample size, restricted geographic representation, and narrow focus on ASTs and detecting uropathogens for MDR patterns constrain its applicability to broader populations. Additionally, certain potential confounders, including patients’ nutritional status, co-morbidities, socioeconomic factors, antibiotic regimens and precise antibiotic usage history, were not accounted for due to insufficient data, potentially influencing the interpretation of MDR determinants.

Despite assessing seasonality, the study was unable to establish a direct association between seasonal variations and MDR-UTIs, highlighting limitations in integrating environmental data and controlling for confounding climatic factors. Moreover, the cross-sectional design inherently restricts cause-and-effect relationships between the identified predictors and MDR status. Therefore, longitudinal studies are recommended further to establish a strong comprehensive understanding of the association between the predictors and risk factors for causing MDR in UTI patients.

### Recommendations

A comprehensive approach is essential to mitigate the spread and impact of MDR pathogens. Robust AMS programs should enforce stringent antibiotic regulations, enhance public awareness, and promote evidence-based prescribing. Infection control must prioritize high-risk settings through improved hygiene, disinfection, and reduced invasive procedures. Pathogen-specific treatment protocols tailored to resistance patterns, continuous surveillance using advanced molecular tools, and investments in healthcare infrastructure are imperative. Expanding public education on antibiotic use and fostering interdisciplinary research through one health approach will further strengthen efforts to combat MDR-UTIs globally.

In conclusion, our study highlights the multifaceted predictors and risk factors, including healthcare-associated factors, patient demographics, and bacterial isolates, with females exhibiting a higher prevalence of UTIs but no direct relationship to MDR status. Healthcare environments, particularly specialized units and private medical settings, emerged as critical contributors, underscoring the need for targeted interventions. The disproportionate impact of pathogens, especially *Acinetobacter* spp., necessitates pathogen-specific strategies. Future studies are suggested to identify the factors or causes contributing to the high MDR rates in SUs and PMSs, despite the OPD handling a substantial volume of cases and treatments.

## Data Availability

Data can be made available upon reasonable request to

## Acknowledgments

We sincerely acknowledge the guidance and ethical oversight provided by the Institutional Review Board (IRB) of Sher-E-Bangla Medical College throughout this study. Additionally, we extend our gratitude to the laboratory staff at the Department of Microbiology, Sher-E-Bangla Medical College, Barishal, for their valuable assistance with sample processing and data collection.

## Author contributions

**Conceptualization** Ibrahim Khalil

**Formal Analysis** Ibrahim Khalil

**Investigation** Ibrahim Khalil, A.K.M. Akbar Kabir

**Methodology** Ibrahim Khalil, Abu Sayed, A.K.M. Akbar Kabir

**Resources** A.K.M. Akbar Kabir, Ibrahim Khalil, Abu Sayed, S. M. Iqbal Hossain

**Software** Ibrahim Khalil, Abu Sayed

**Supervision** Abu Sayed, A.K.M. Akbar Kabir

**Visualization** Ibrahim Khalil, Abu Sayed

**Writing** – Original Draft Preparation Ibrahim Khalil, Abu Sayed, A.K.M. Akbar Kabir, Md. Nurul Alam, Rahima Akther Dipa, Md. Tanvir Rahman

**Writing – Review & Editing** Abu Sayed

## Use of Artificial Intelligence (AI)-Assisted Technology for manuscript preparation

During the preparation of this manuscript, the authors utilized ChatGPT (version 4.0) for linguistic improvements. The authors carefully reviewed and revised the content to ensure accuracy, completeness, and adherence to the journal’s standards and take full responsibility for the final content of the published article.

